# HLA Genotype is Associated with Patient Reported Outcomes and Adverse Reactions to Metal Debris in Patients with Cobalt-Chrome Knee Arthroplasties

**DOI:** 10.64898/2026.10.12.26365276

**Authors:** David J Langton, Rohan Bhalekar, Yash Borikar, Stephen Wells, Nick Clement, David Deehan, Dominie Guest, Chris Harrington, Simon Jameson, Gillian Leitch, Rachael McManus, Moreica Pabbruwe, Jon Phillips, Andrew Porteous, Steven Rushton, Alasdair Santini, Samantha Tross, Raghavendra Sidaginamale

## Abstract

**Purpose:** To determine whether a cobalt-chrome genetic reactivity (CoGR) score - originally developed to identify patients at greater risk of developing adverse reaction to metal-on-metal hips - is associated with poorer patient-reported outcomes after knee arthroplasty (KA).

**Methods:** In a prospective multicentre study, patients at a minimum two years post all cobalt-chrome KA were invited to research clinics. Participants provided samples for blood metal ion measurements and DNA sequencing and completed Forgotten Joint Scores (FJS). Component alignment was assessed on AP and lateral plain radiographs.

The association between CoGR and FJS was assessed using regression adjusted for time since surgery, age, gender, BMI and tested for modification by blood cobalt. In a case-control analysis, CoGR scores of revised patients with histologically confirmed adverse reactions to metal debris (and specifically aseptic lymphocyte-dominated vasculitis-associated lesions (ALVAL)) were compared with controls (revised patients with no signs of ARMD and unrevised patients with FJS of 75 or more) using Firth logistic regression.

**Results:** 376 patients were included: 312 unrevised patients and 64 patients who underwent revision. Each one-unit increase in CoGR score was associated with a 9.8 point lower FJS (95% confidence interval (CI) 4.2 to 15.4 points; p < 0.001). This increased to a 17.5 (9.4 to 25.5) point reduction per CoGR score unit in patients with blood cobalt above the median level. Comparing 28 ARMD cases (12 with ALVAL) with 177 controls, each one-unit CoGR increase was associated with odds ratios of 1.90 (CI 1.34 to 2.75) for ARMD and 3.05 (CI 1.89 to 5.34) for ALVAL.

**Conclusions:** A higher CoGR score was associated with worse patient-reported outcome and higher odds of histologically confirmed adverse reaction to metal debris at revision. Adverse immune responses may be a more significant factor in unexplained pain after KA than is generally recognised.

## Introduction

Total knee arthroplasty (TKA) reduces pain and improves quality of life in the majority of patients.[1] However, a significant minority develop persistent pain or dissatisfaction, despite efforts to improve outcomes that have largely focused on optimising implant design and component alignment.[2] Less attention has been paid to immunogenetic factors. Although the vast majority of TKAs incorporate at least one cobalt-chromium (CoCr) alloy component[3], the influence of the host immune response to CoCr on patient reported outcome measures (PROMs) remains poorly understood.[4]

Adverse reactions to metal debris (ARMD), including aseptic lymphocyte-dominated vasculitis-associated lesion (ALVAL), are well described in – yet not restricted to – patients with metal-on-metal hip arthroplasties.[5, 6] We have previously shown that the development of ARMD following metal-on-metal hip arthroplasty is determined by an interaction between the dose and duration of metal exposure and specific genetic variants.[7] We subsequently developed the cobalt-chrome alloy genetic reactivity (CoGR) score to quantify an individual’s genetic predisposition to CoCr driven ARMD. The CoGR score forms the basis of the commercially available test Orthotype™ (ExplantLab, Newcastle Upon Tyne, United Kingdom), which was developed by some of the authors.

The CoGR score uses the output of next-generation DNA sequencing to reconstruct a patient’s human leucocyte antigen (HLA) encoded major histocompatibility complexes (MHCs). Based on insilico electrostatic and peptide binding calculations, each patient HLA genotype is assigned a risk score; a higher CoGR score indicates a greater genetic predisposition to developing ARMD and ALVAL when exposed to equivalent amounts of CoCr metal debris.

We hypothesised that if genetically determined reactivity to CoCr contributes to symptoms below the threshold of revision, it may account for a proportion of otherwise unexplained dissatisfaction following TKA. The primary aim of this study, therefore, was to determine whether higher CoGR scores were associated with lower PROMs following all CoCr TKA. As a secondary aim, we examined whether higher CoGR scores are associated with an increased incidence of histologically confirmed ARMD and ALVAL in patients undergoing revision.

## Methods

This study was performed within an ongoing prospective, multicentre investigation of the influence of blood metal ion concentrations and patient genetics on the outcome of arthroplasty. The overarching study was not limited to any specific device, joint or materials. However, the current paper includes only those patients with all-CoCr knee arthroplasties (femoral and tibial components both manufactured from CoCr alloy) (Fig. 1), in order to isolate metal exposure to CoCr alloy only.

**Fig. 1.**
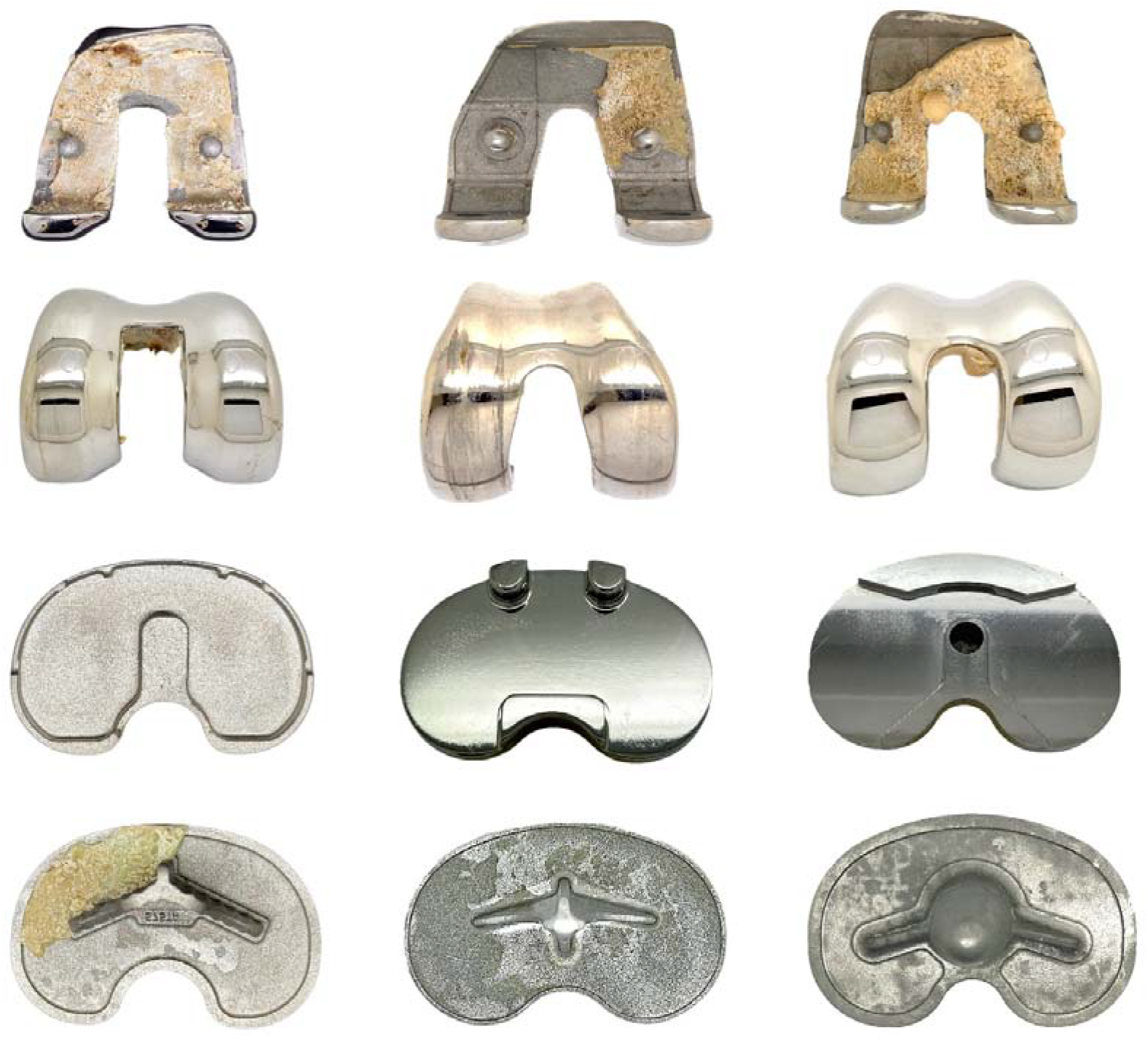
The most common arthroplasties implanted into the patients in this study were Triathlons (Stryker, Kalamazoo, Michigan) (left), Vanguard (Zimmer Biomet, Warsaw, Indiana) (middle) and Attunes (Depuy Synthes, Warsaw, Indiana) (right). Explanted prostheses are shown, with all CoCr surfaces visible.

Two patient groups were recruited: patients a minimum of two years post arthroplasty (“unrevised patients”), and patients undergoing revision arthroplasty (“revision patients”).

At participating centres, unrevised patients were identified through local hospital records and invited to a review clinic or recruited while attending for review/consideration of other joint surgery. All consenting patients undergoing revision surgery for any cause at the centres were included. Consenting patients completed PROMs questionnaires (the Forgotten Joint Score (FJS)) and gave blood for next-generation DNA sequencing and measurement of blood metal ion concentrations. DNA extraction, sequencing and metal ion measurements were carried out as previously described.[7] All available anteroposterior and lateral plain radiographs were reviewed and basic measures of component alignment recorded. These included: the lateral distal femoral angle (LDFA); the medial proximal tibial angle (MPTA) and joint line obliquity (JLO).[8] Neither patients attending clinics nor medical staff overseeing the follow ups, carrying out revisions or performing histological reviews were aware of the results of the DNA sequencing or blood metal ion concentrations.

### Part 1: Unrevised patients

In the unrevised patients we tested two hypotheses:

1. Higher CoGR grades would be associated with lower FJS
2. The influence of CoGR on FJS would be greater in patients with higher blood metal ion concentrations

### Are higher CoGR grades associated with lower FJS?

The FJS extends from 0 to 100. It is recognised that many patients reach the top of the scale, and a lower proportion, the bottom. A patient scoring 100 may be doing even better than the scale can show, and ordinary regression treats such scores as exact measurements, which tends to understate real associations. We therefore used Tobit regression, which allows for scores being capped at both ends by creating a latent variable whose range can exceed the caps at upper and lower limits. The analysis then estimates the association on the underlying, uncapped score [9, 10]. Results are expressed as points on that underlying score and are somewhat larger than ordinary regression estimates, which we also report as a check. CoGR grade (a whole number from −2 to 4; higher grades mean greater genetic reactivity to CoCr) was treated as an explanatory variable along with other factors that might influence FJS: duration since implantation, component alignment, age at surgery, body mass index (BMI), gender and blood metal ion concentrations. A ‘full’ model with all covariates included was considered to assess relative contributions of each covariate.[11, 12] Because unrevised patients were assessed at a single review at least two years after surgery rather than followed from implantation, time-to-event methods such as Cox regression do not apply; time since implantation was therefore included as a covariate to assess the extent to which duration of exposure to the implant might impact on the outcome. Blood metal concentrations were log transformed due to their non parametric distribution (Appendix A).

### Is the influence of CoGR on FJS greater in patients with higher metal ion concentrations?

If CoGR identifies patients who react to metal debris, its association with FJS should be stronger when metal exposure is higher. We therefore compared the CoGR–FJS association in patients with blood cobalt (Co) above and below the median concentration. Because a single cut-point is arbitrary and may have influenced the model outcomes, we repeated the analysis using tertiles and quartiles of the Co distribution and with Co as a continuous measure. Appendix Fig. B1 shows the distribution of Co with these thresholds marked.

#### Model checks and software

We checked that the model assumptions were reasonable and that no single patient was driving the findings (Appendix A). Results are shown with 95% confidence intervals (CI). Two-sided p < 0.05 was considered statistically significant. Analyses used Python 3.12 (pandas, SciPy, statsmodels) and XLSTAT.

### Part 2: Is CoGR score associated with an increased risk of ARMD and ALVAL: Case–Control Analysis

“Cases” were defined as revised patients with histologically confirmed ARMD.[13] “Controls” were defined as patients judged free of clinically significant ARMD/ALVAL on one of two grounds: unrevised patients with a FJS ≥ 75, or revised patients whose histology was negative for ARMD. Revised patients with no tissue sent for histology and unrevised patients with FJS < 75 were excluded, as their disease status could not be established.

We compared CoGR between cases and controls and expressed the difference as an odds ratio per one-grade increase in CoGR. For example, an odds ratio of 2 means that the odds of disease double with each grade. With relatively low numbers of cases, standard logistic regression is known to overestimate associations, so Firth’s penalised logistic regression was used, which corrects this to some extent [14, 15, 16]. It cannot add information that is not in the data, so confidence intervals remain wide when case numbers are low, and it has a small bias of its own in predicted probabilities [17]. We therefore repeated every analysis with two published refinements (Firth logistic regression with intercept correction, FLIC, and with added covariate, FLAC) (Appendix A). As checks that do not depend on a model, we compared the proportion of patients with CoGR ≥ 1 between groups (Fisher’s exact test) and the distribution of CoGR grades (Mann–Whitney U test). To test specificity, we compared revised patients with and without ARMD.

A large language model was used to optimise manuscript layout, language and check statistical outputs.

## Results

### Patients

In total, there were 376 patients with an all-CoCr knee arthroplasty: 312 unrevised patients, assessed a median of 9.0 years after surgery (interquartile range, IQR, 7.0–11.0), and 64 patients undergoing revision. Recruitment rate varied between 40 and 50% of eligible patients across the hospitals for both revised and unrevised patients. Age was missing for 6 patients and BMI for 10 (under 4% in each case). These patients were omitted from models using those variables (complete-case analysis, n = 296), which was unlikely to bias results as so few data were missing (Appendix A). Among the unrevised patients, 162 (51.9%) were female, the mean age at primary surgery was 64.1 years (standard deviation, SD, 8.0) and mean BMI was 32.4 kg/m^2^ (SD 6.7). Median FJS was 75.0 (IQR 31.7–97.7), with 22.1% of patients reporting the maximum score of 100 and 4.5% the minimum of 0, confirming the need for an analysis that allowed for capped scores (Appendix Fig. B1). Median CoGR grade was 0 (IQR 0–1). Median blood Co was 0.35 µg/L (IQR 0.22–0.71) and median blood chromium (Cr) 0.48 µg/L (IQR 0.35–0.78). Radiographs were available for 260 of the 312 unrevised patients.

For the case–control analysis, 28 revised patients had histologically confirmed ARMD (12 of whom had ALVAL) and 177 were controls: 158 unrevised patients with FJS ≥ 75 and 19 revised patients whose histology was negative for ARMD. Seventeen revised patients with no tissue sent for histology, and 154 unrevised patients with FJS < 75, were excluded because their disease status could not be established. Table 1 provides details of the patient groups.

**Table 1.** Characteristics of each patient group for the case control analysis.

| | Controls —<br>unrevised, FJS $\geq 75$ (n<br>= 158) | Controls —<br>revised,<br>histology<br>negative (n = 19) | ARMD (n = 28) | ALVAL subset (n<br>= 12) | p* |
| --- | --- | --- | --- | --- | --- |
| Age at primary (yrs) | 66 [60–71] | 60 [56–66] | 58.5 [52–66] | 63.5 [58–66] | 0.002 |
| Male | 80 (51%) | 10 (53%) | 12 (43%) | 5 (42%) | 0.54 |
| BMI (kg/m<sup>2</sup>) | 30.4 [26.5–34.0] | 33.1 [30.6–37.0] | 30.9 [26.3–35.1] | 31.0 [29.9–35.5] | 0.99 |
| Duration in vivo (yrs) | 10 [8–11] | 7 [1.3–14] | 11.5 [5.8–17] | 9.5 [3.1–16] | 0.17 |
| Blood Co (µg/L) | 0.36 [0.23–0.75] | 0.48 [0.22–0.64] | 3.10 [0.46–20.5] | 7.36 [0.52–20.8] | < 0.001 |
| Blood Cr (µg/L) | 0.46 [0.35–0.71] | 0.56 [0.50–0.78] | 1.62 [0.60–4.98] | 2.98 [0.60–7.05] | < 0.001 |

\*p compares ARMD cases with all 177 controls (Mann–Whitney or Fisher)

### Part 1: Unrevised patients

#### Is a higher CoGR grade associated with a lower FJS?

Patients with higher CoGR grades reported lower FJS (Fig. 2). After adjustment for time since surgery, age, BMI, gender and blood metal ion concentrations, each one-grade increase in CoGR was associated with a 9.8-point lower FJS on the underlying scale (95% CI 4.2 to 15.4 points lower; p < 0.001; Table 2). In simplistic terms, a patient with a CoGR grade of 3 would be expected to score around 29 points lower than an otherwise similar patient with grade 0. Each grade had an effect similar in size to a 10-unit (kg/m^2^) difference in BMI. Ordinary regression, which ignores the capped scores, gave a smaller but significant estimate (7.0 points lower per grade, p = 0.003).

**Table 2.**
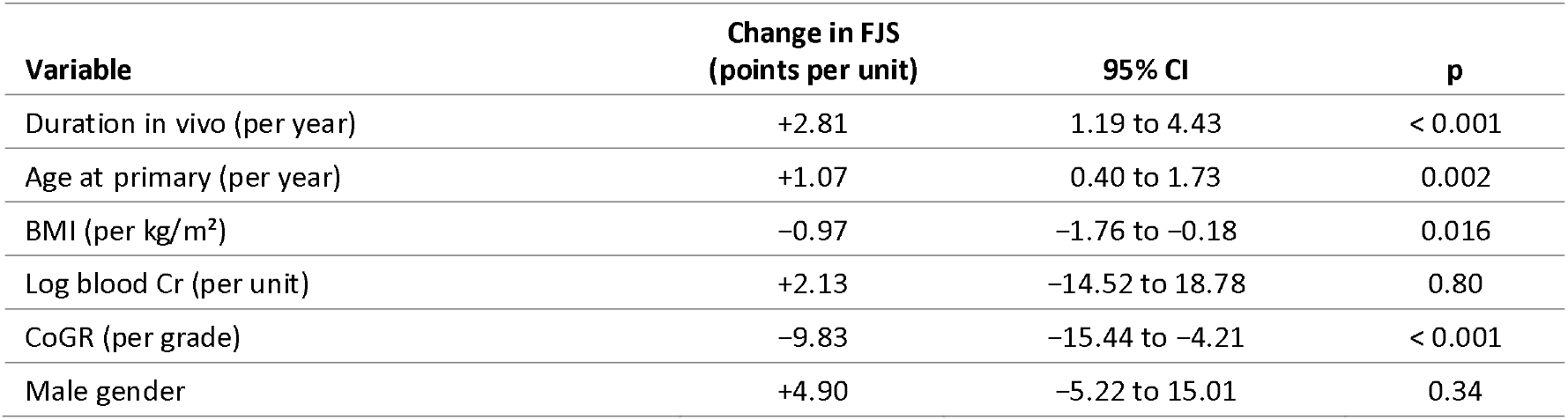
Factors associated with Forgotten Joint Score in unrevised patients (two-limit Tobit regression, n = 296 complete cases; pseudo-R2 = 0.14). Coefficients are points on the underlying FJS scale per one-unit increase. No measured radiographic parameters significantly impacted the modelling (Appendix C).

| Variable | Change in FJS<br>(points per unit) | 95% CI | p |
| --- | --- | --- | --- |
| Duration in vivo (per year) | +2.81 | 1.19 to 4.43 | < 0.001 |
| Age at primary (per year) | +1.07 | 0.40 to 1.73 | 0.002 |
| BMI (per kg/m<sup>2</sup>) | -0.97 | -1.76 to -0.18 | 0.016 |
| Log blood Cr (per unit) | +2.13 | -14.52 to 18.78 | 0.80 |
| CoGR (per grade) | -9.83 | -15.44 to -4.21 | < 0.001 |
| Male gender | +4.90 | -5.22 to 15.01 | 0.34 |

**Fig. 2.**
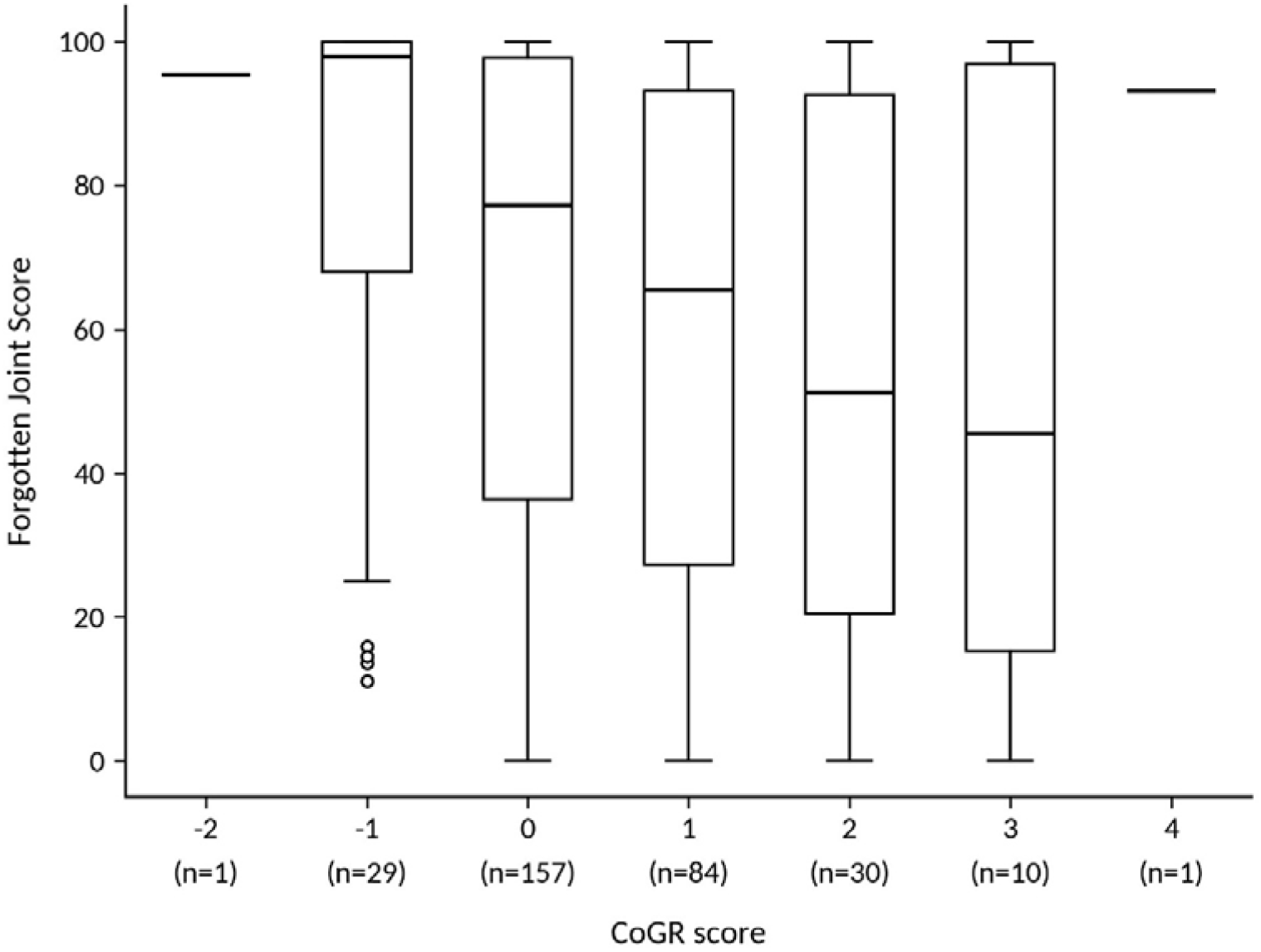
The distribution of FJS in each CoGR grade patient group. Horizontal lines represent the median values in each group with the boxes representing the upper and lower quartiles. The whiskers represent the limits of the data with circles the outliers.

Three other factors were independently associated with FJS (Table 2). Scores were higher with longer time since surgery (+2.8 points per year, p < 0.001) and older age at surgery (+1.1 points per year, p = 0.002), and lower with higher BMI (−1.0 point per kg/m^2^, p = 0.016). Gender and blood Cr (or Co) were not associated with FJS, and neither were radiographic measures of alignment (all p ≥ 0.39; Appendix Table C1). A more complete description of the radiographic measures is included in Appendix C.

#### Is the association between CoGR and FJS stronger when blood Co is higher?

In patients with blood Co above the median (n = 141), each grade of CoGR was associated with a 17.5-point lower FJS (95% CI −25.5 to −9.4; p < 0.001). In patients at or below the median (n = 155) the association was weaker and did not reach statistical significance (−4.4 points; 95% CI −12.0 to 3.2; p = 0.25). The result did not depend on the choice of ion threshold. The effect of CoGR increased steadily across tertiles of the Co distribution, from −4.9 points per grade in the lowest tertile, to −6.7 in the middle tertile and −19.4 in the highest tertile (95% CI −28.9 to −9.9; Fig. 3), and treating Co as a continuous measure gave the same conclusion (interaction p = 0.022). Analysis using quartiles also produced the same pattern (Appendix Table B4).

**Fig. 3.**
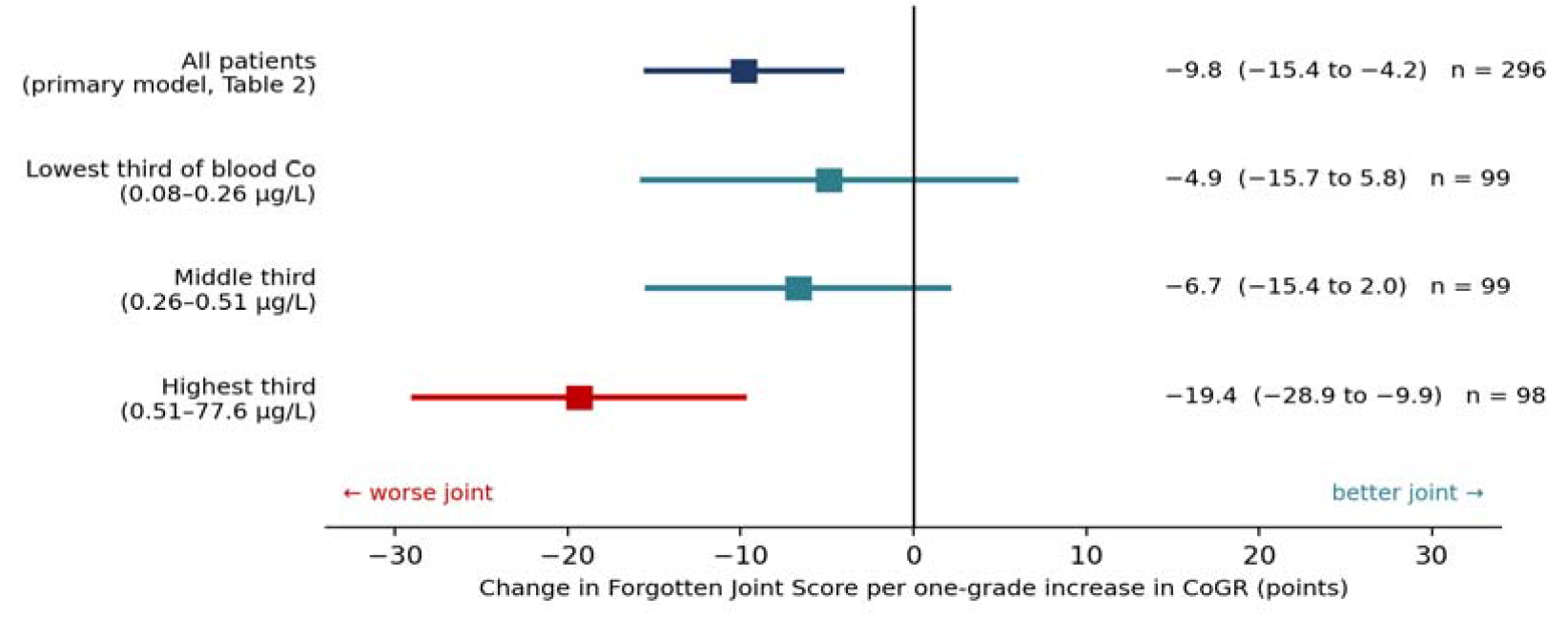
Change in Forgotten Joint Score (FJS) per one-grade increase in CoGR, overall and within tertiles of the distribution of blood Co concentration. Points are estimates and bars 95% confidence intervals from Tobit models. The effect of CoGR was largest in patients with the highest Co concentrations.

Removing the two non-significant adjustment factors (blood Cr and gender) left the CoGR estimate essentially unchanged (−10.2 vs −9.8 points per grade; Appendix Table B2). Grouping CoGR into <0, 0, 1 and ≥2 rather than treating it as a straight line gave the same stepwise pattern (Appendix B.5). The model assumptions were satisfactory, the predictors were not meaningfully overlapping (variance inflation factors below 1.2), and no single patient changed the CoGR estimate by more than one point (Appendix B.3).

##### Part 2: Are CoGR scores higher in patients with histologically confirmed ARMD?

CoGR scores were significantly higher in patients with ARMD, and the association strengthened with the severity of the histological change. A CoGR grade of 1 or more was present in 34% of unrevised controls (53 of 158), 11% of revised patients without ARMD (2 of 19), 68% of patients with ARMD (19 of 28) and 92% of those with ALVAL (11 of 12; Appendix Table B5). Fig. 4 shows how often ARMD and ALVAL were found at each CoGR grade: ARMD rose from 4–5% in patients with grade 0 or below to 19% at grade 3 or above, and ALVAL from 0–1% to 19%.

**Fig. 4.**
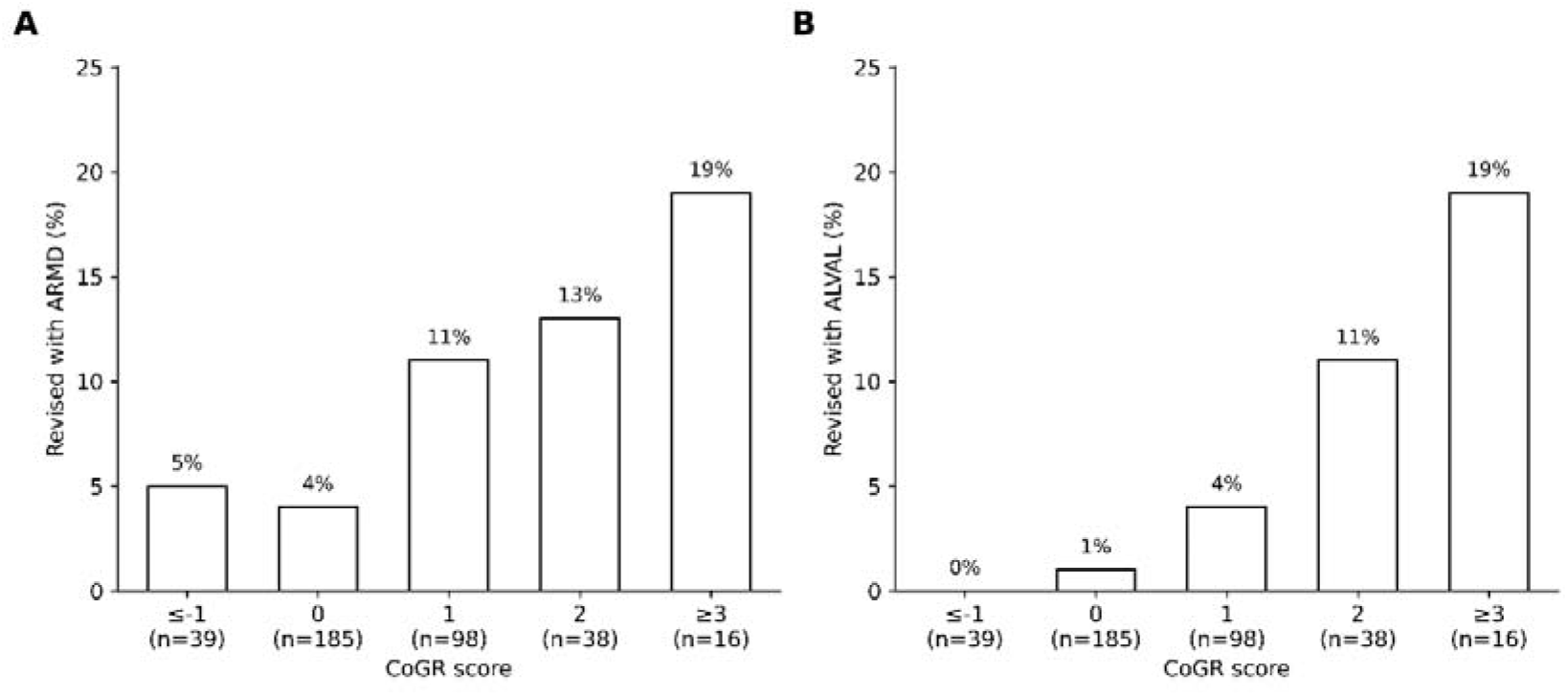
**A.** Percentage of patients in each CoGR group with histological signs of ARMD. **B**. Percentage of patients in each CoGR group who were found to have histological signs of ALVAL.

In regression analysis (Table 3), each one-grade increase in CoGR was associated with approximately double the odds of ARMD (odds ratio 1.90; 95% CI 1.34–2.75; p < 0.001) and triple the odds of ALVAL (odds ratio 3.05; 95% CI 1.89–5.34; p < 0.001). Adjustment for age, BMI, gender and time since implantation (ARMD), or for age and gender (ALVAL, given the small number of cases), made no material difference (ARMD 1.97; ALVAL 3.12). Younger age at primary surgery was the only other factor independently associated with ARMD (odds ratio 0.93 per year; 95% CI 0.88–0.97; p = 0.001). There were only 12 cases of ALVAL, therefore confidence intervals were wide.

**Table 3.**
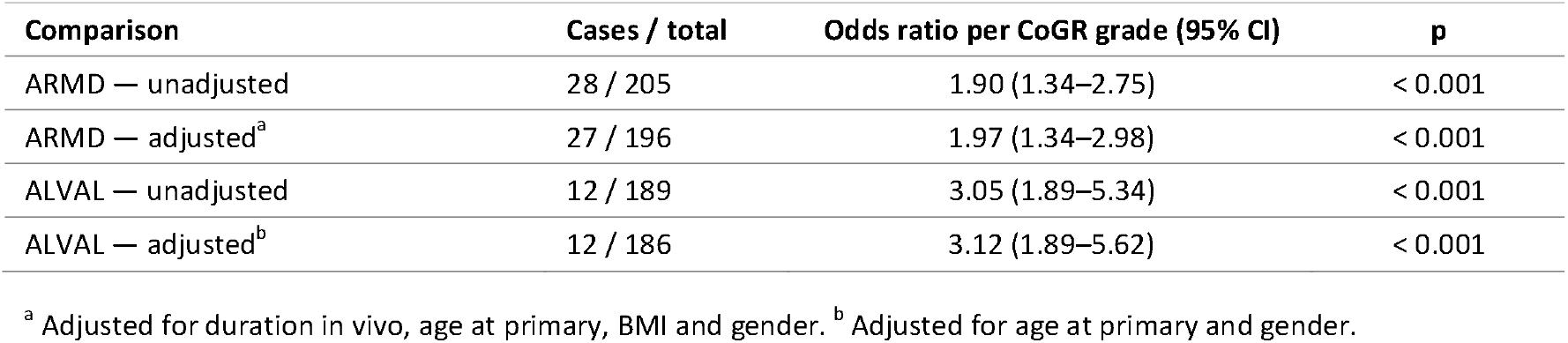
Odds of disease per one-grade increase in CoGR (Firth logistic regression).

A model-free comparison gave consistent results. A CoGR grade of 1 or more was present in 68% of ARMD cases and 31% of controls (Fisher’s exact odds ratio 4.68; p < 0.001). The finding was specific to ARMD rather than to revision in general. Only 2 of 19 revised patients without ARMD had a grade of 1 or more, compared with 19 of 28 patients with ARMD (p < 0.001). Their CoGR grades were significantly lower than those of well-functioning unrevised controls (p = 0.040).

Odds ratios were similar between the standard logistic regression and the Firth, FLIC and FLAC methods (ARMD 1.88–2.05; ALVAL 2.97–3.39; Appendix Table B6), and there was no sign that the data were separated. There was weak evidence that the rise in odds flattened at the very highest CoGR grades, where numbers were small (3–5 patients at grades 3 and 4).

## Discussion

The principal finding of this study is that a genetic risk score for ARMD developed through the study of patients with metal-on-metal (MOM) hips - the CoGR score - is associated with reduced functional outcome in patients with CoCr knee arthroplasties. In this large cohort of unrevised patients, each CoGR grade was associated with a 9.8-point lower FJS. This association was concentrated in patients with blood Co concentrations above the cohort median, in whom the effect reached 17.5 points per grade. In terms of confirmed histopathological response, each one-grade increase in CoGR approximately doubled the odds of ARMD and tripled the odds of ALVAL. These estimates were essentially unchanged after adjustment for age, gender, BMI and duration in vivo and the associations held across multiple analytic approaches. Cases with ARMD had markedly higher blood Co and Cr and higher CoGR scores than controls, with the highest levels in the ALVAL subset, consistent with a gene–environment interaction underlying a progressive spectrum of disease.

### Associations with lower functional outcomes and investigation of painful knees

It is recognised that 10-20% of patients report pain, stiffness or dissatisfaction after knee arthroplasty.[18] Efforts to address dissatisfaction after knee arthroplasty have largely concentrated on variables within surgical control, such as improving the precision of component alignment as well as changes to prosthetic constraint[19] and alignment philosophy.[20] Relatively little attention is paid to the choice of implant material, with adverse immune responses to metal given little significance in patient outcomes.[21] Protocols for the investigation of painful arthroplasties reflect this. The “painful prosthesis” is frequently defined as persistent pain in the absence of infection, loosening or malalignment.[18] Hypersensitivity to metal, where acknowledged at all, is listed among unusual causes rather than being viewed as a true diagnostic category with an accepted workup. Crucially, there are no widely used tests to diagnose adverse immune responses in patients with knee arthroplasties.

### Problem with diagnostics

Where metal sensitivity/allergy is considered at all, attention has tended to focus on nickel sensitivity and skin patch testing. Patch testing has well-documented weaknesses, including poor agreement between testing modalities and an uncertain relationship between patch-test positivity and clinical outcome.[22, 23] Another testing modality is the lymphocyte transformation test (LTT).

LTT was adapted for orthopaedic application at approximately the same time as — and independently of — the first full histological characterisation of ALVAL in 2005.[24] The assay therefore predates the disease entity it is now used to detect. Its diagnostic performance, when assessed against histologically proven ARMD or ALVAL has, to our knowledge, not been established in large case numbers.[6, 25] A further concern is that both LTT and skin patch testing protocols involve the exposure of cells or skin to differing concentrations of metals which often greatly exceed those typically seen in the periprosthetic environment.[26]

### Gold standard and development of knowledge

Dermatologists have not reached consensus about whether ALVAL is a histological manifestation of metal hypersensitivity/allergy.[27] However, in orthopaedics, the clinical relevance of ALVAL has been established. It is associated with greater levels of pain in patients with hip and knee arthroplasties, increased synovial thickness on magnetic resonance imaging (MRI) and the extent of tissue destruction at revision.[28, 29] There is emerging evidence that ARMD may be under-recognised in knee arthroplasty, with signs of ALVAL reported in approximately 30% of tissue samples taken from patients undergoing revision.[5, 30, 31] There is also evidence that ALVAL may masquerade as culture negative prosthetic joint infections, just as was the case with the first metal-on-metal ALVAL reactions encountered by unsuspecting surgeons.[31, 32]

The history of metal-on-metal hip arthroplasty shows other parallels: early metal reactions were viewed with scepticism and ascribed to nickel sensitivity in unusually susceptible individuals[33]; patch testing and LTT were initially recommended but proved unreliable[6, 25]; and the clinical use of blood metal ion testing was questioned for years before becoming routine in follow-up clinics across the world.[34, 35] Even after ALVAL was widely recognised, pain after metal-on-metal arthroplasty was still being attributed to surgical causes.[7]

While we believe delayed type metal allergic responses are more important than is generally recognised, we do not believe that ALVAL develops in all genetically susceptible people undergoing knee arthroplasty (KA). Evidence from this study, and previous hip and knee studies, appear to be in unison that an ALVAL response generally requires marked elevations in metal ion exposure to become fully established.[36, 37, 38] This implies that most individuals require sustained exposure to large local concentrations of metal debris before tolerance is broken, a more aggressive inflammatory response is mounted, and bone and/or soft tissue are put at risk. The typical volume of metal debris released from a well-functioning KA does not result in sufficient metal ion exposure to breach that threshold in the vast majority of patients (Table 1).

We suspect, however, that genetically susceptible individuals mount a milder, macrophage-driven response at lower levels of metal exposure, resulting in vague symptoms with less severe microscopic and macroscopic findings. By definition, the histopathological response in these unrevised patients cannot be determined. We therefore suggest the term genetically associated arthroplasty pain, histologically ill-defined (GEN-APHID). As GEN-APHID, ARMD and ALVAL share the same genetic risk (Figs 3 and 4), this implies they form a spectrum of the same disease process, consistent with the graded CoGR-ARMD–ALVAL relationship we have reported.

### Limitations

We emphasise that we do not believe that all the symptomatic patients in the study were suffering from metal related pathology. We also stress that the findings of this paper do not diminish the importance of other established contributors to outcome, including implant selection, surgical technique, patient selection, psychological factors and other variables we did not quantify.[39, 40] The main limitation affecting the study is potential selection bias. The combined case–control analysis drew cases and controls from different clinical settings, and control status was defined by a FJS of 75 or greater rather than by histology. While we believe this definition is reasonable, occult ARMD among controls cannot be excluded, though such misclassification would be expected to reduce the strength of associations. Event counts were modest, particularly for ALVAL (n = 12), and although Firth penalisation mitigates small-sample bias, the confidence intervals around the ALVAL estimates were wide. In addition, the functional outcome analysis was cross-sectional, precluding assessment of clinical trajectory.

## Conclusion

In patients with all-CoCr knee arthroplasty, the CoGR score was associated with worse patient-reported outcomes in a metal ion concentration-dependent manner, and with histologically confirmed ARMD and ALVAL at revision in a dose-dependent fashion. These findings suggest that ARMD is not a phenomenon confined to metal-on-metal hip arthroplasty, but may be an important under-recognised contributor to unexplained pain after knee arthroplasty.

## Abbreviations

ALVAL: aseptic lymphocyte-dominated vasculitis-associated lesion
ARMD: adverse reaction to metal debris
BMI: body mass index
CI: confidence interval
Co: cobalt
CoCr: cobalt-chromium
CoGR: cobalt-chrome alloy genetic reactivity
Cr: chromium
FJS: Forgotten Joint Score
FLAC: Firth logistic regression with added covariate
FLIC: Firth logistic regression with intercept correction
GEN-APHID: genetically associated arthroplasty pain, histologically ill-defined
HLA: human leucocyte antigen
IQR: interquartile range
JLO: joint line obliquity
KA: knee arthroplasty
LDFA: lateral distal femoral angle
LTT: lymphocyte transformation test
MHC: major histocompatibility complex
MOM: metal-on-metal
MPTA: medial proximal tibial angle
MRI: magnetic resonance imaging
PROMs: patient reported outcome measures
SD: standard deviation
TKA: total knee arthroplasty

## Declarations

### Ethical approval

The study was conducted under IRAS (Integrated Research Application System) project 14119 (REC reference 09/H0905/41). REC approval was obtained from the County Durham and Tees 2 board.

### Informed consent

Informed written consent was obtained from all individual participants included in the study.

### Data availability

Individual participant data, including genetic data, are not publicly available because of participant privacy and the terms of consent. All data supporting the findings are summarised in the manuscript and its Appendix. Requests for further information may be directed to the corresponding author.

### Reporting guideline

STROBE guidelines were followed.

### Use of artificial intelligence tools

A large language model was used to assist with manuscript layout and language editing, and to help implement and check the statistical analyses. All analyses, results and text were reviewed and verified by the authors, who take full responsibility for the content.

## Appendix (supplementary material)

### Appendix A. Statistical methods in more detail

#### A.1 Tobit regression

FJS is bounded at 0 and 100. When a substantial proportion of patients score at either bound, ordinary least squares (OLS) regression treats these bounded observations as precise, unbounded measurements, which attenuates estimated associations and can bias inference, particularly among higher-functioning patients. This was anticipated given the well-described ceiling effect of FJS and confirmed in this cohort (22.1% at 100; 4.5% at 0; Appendix Fig. B1). A two-limit Tobit model, censored at 0 and 100, was therefore the primary multivariable analysis [9, 10]. It assumes an underlying (latent) continuous score, normally distributed around a linear function of the predictors, of which only the part between 0 and 100 is observed; coefficients are effects on this latent score and were estimated by maximum likelihood. Univariable screening used rank-based methods (Spearman correlation, Mann–Whitney U test), which assume no particular distribution, because FJS is not normally distributed (Shapiro–Wilk p < 0.001). The multivariable analysis necessarily moves to a parametric framework because no rank-based method adjusts for several covariates while allowing for censoring at both bounds. An OLS model was fitted in parallel as a sensitivity analysis. Model performance was summarised by pseudo-R^2^.

#### A.2 Transformation, variable selection and missing data

Blood Co and Cr concentrations were log-transformed because both were strongly right-skewed in the unrevised cohort (skewness 10.8 for Co and 4.3 for Cr, falling to 1.6 and 1.5 after transformation) and Co spanned approximately three orders of magnitude (0.08–77.6 µg/L), so untransformed values would be dominated by a few extreme observations. All candidate variables were entered into the multivariable model simultaneously, irrespective of univariable significance, because selecting variables on univariable p-values is known to give biased and unstable models and can discard variables that act jointly with others [11, 12]. Age was missing for 6 patients (1.9%) and BMI for 10 (3.2%), 16 patients in total.

These patients did not differ from the rest in FJS (median 74.0 vs 75.0, p = 0.70) or CoGR (p = 0.51), so complete-case analysis (n = 296) was judged unlikely to introduce meaningful bias and was preferred over multiple imputation to avoid the additional distributional assumptions the latter would require.

#### A.3 Effect modification by cobalt

High cobalt was defined as above the median blood Co of the 296 complete cases (0.34 µg/L). Two analyses were used: an interaction Tobit model including a high-cobalt indicator and its product with CoGR, tested with the likelihood-ratio test; and Tobit models fitted separately within each stratum. Both were adjusted for duration in vivo, age at primary, BMI and gender. Because a median is an arbitrary cut-point, the models were repeated within tertiles (boundaries 0.26 and 0.51 µg/L) and quartiles (0.22, 0.34 and 0.70 µg/L), the interaction between CoGR and continuous log Co was tested, and log Cr was added to the interaction model.

#### A.4 Firth regression, FLIC and FLAC

Conventional maximum-likelihood logistic regression is biased away from the null when events are few, and its estimates can fail to exist (becoming infinite) when a covariate pattern perfectly separates cases from controls. Firth’s method adds a penalty to the likelihood (equivalent to a Jeffreys prior) that removes the leading term of this small-sample bias and always returns finite estimates [14, 15]. It was chosen because event numbers were small (28 ARMD and 12 ALVAL cases), giving only 5.4 and 4.0 events per covariate in the adjusted ARMD and ALVAL models; the traditional rule of ten events per variable is not a fixed threshold [41], but sparse-data bias is a recognised risk below it. Maximum-likelihood estimates were obtainable in every model (no separation), so Firth’s method was used for bias reduction rather than to rescue non-convergent fits. Confidence intervals and p-values were based on the profile penalised likelihood, which does not rely on the normal approximation that is unreliable with few events.

Firth’s method does not guarantee freedom from small-sample problems: it creates no additional information or degrees of freedom, so intervals remain wide when events are few, and it introduces a bias of its own by pulling predicted probabilities towards 0.5, which matters chiefly when predicted risks or marginal effects are estimated from rare events or when separation is present [16, 17]. Regression coefficients, and therefore the odds ratios reported here, are largely unaffected, but to confirm this every model was repeated using two extensions that correct the predicted probabilities: Firth’s logistic regression with intercept correction (FLIC), which re-estimates the intercept so that the mean predicted probability equals the observed event proportion, and with an added covariate (FLAC), which refits the model to data augmented with weighted pseudo-observations and an indicator variable [16]. Odds ratios under maximum likelihood, Firth, FLIC and FLAC are given in Appendix Table B6.

#### A.5 Model checks

Multicollinearity was assessed with variance inflation factors. The Tobit normality assumption was examined with standardised residuals for the non-censored observations (Appendix Fig. B2). Influence was assessed by re-estimating the model omitting each patient in turn. For the logistic models, separation was excluded by confirming finite maximum-likelihood estimates and standard errors, and linearity of the log-odds in CoGR grade was examined by adding a quadratic term (penalised likelihood-ratio test). The model-free comparisons do not depend on these assumptions. FLIC and FLAC were implemented in Python following Puhr et al. [16] and checked against the published worked example of Hashibe et al. [17].

### Appendix B. Additional results

#### B.1 Distribution of FJS and blood cobalt

Appendix Fig. B1 shows the ceiling and floor effects in FJS, the FJS ≥ 75 threshold used to define well-functioning controls, and the distribution of blood cobalt with the cut-points used for stratification.

**Appendix Fig. B1.**
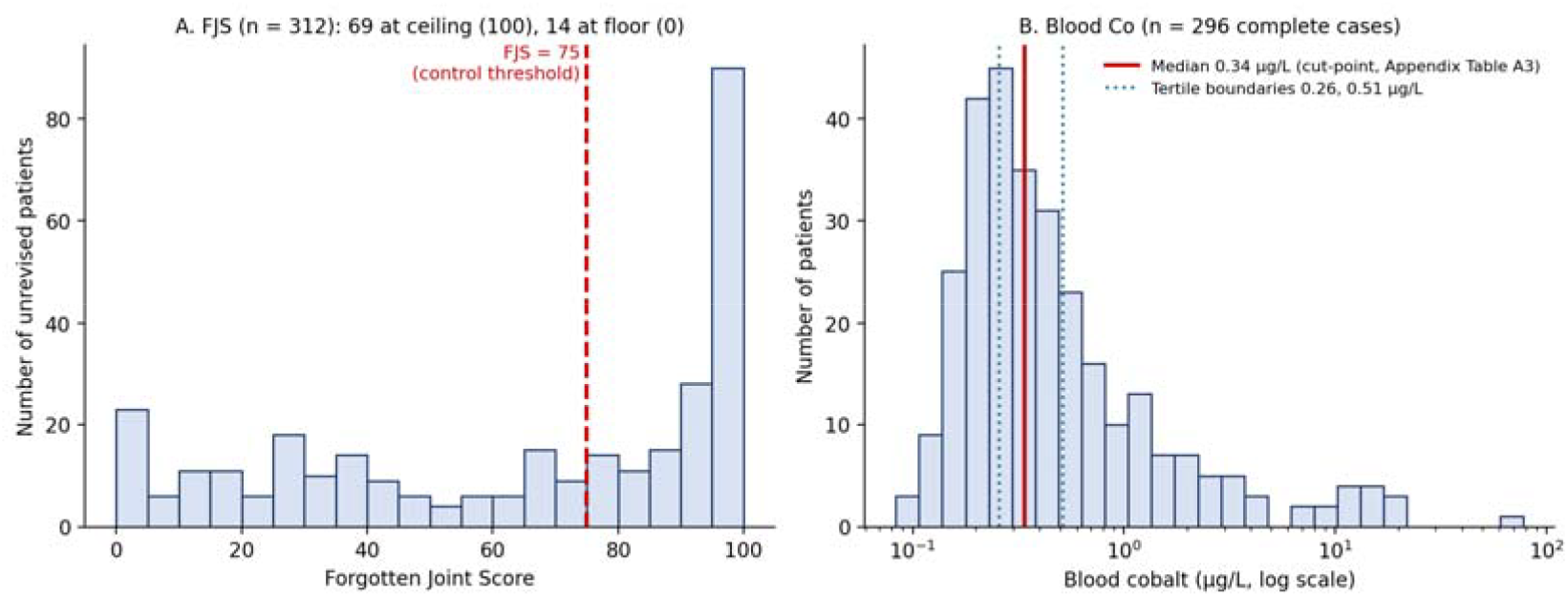
Frequency distributions in unrevised patients. a Forgotten Joint Score, showing the ceiling and floor effects and the FJS ≥ 75 threshold. b Blood Co concentration (log scale), showing the median used as the cut-point in Appendix Table B3 and the tertile boundaries used in Appendix Table B4.

#### B.2 Univariable associations

FJS showed weak but statistically significant positive correlations with duration in vivo, age at primary and log Co, and negative correlations with BMI and CoGR grade (Appendix Table B1). Log Cr was not associated with FJS, and FJS did not differ significantly by gender. There were no significant associations between FJS and radiographic alignment measures (LDFA ρ = 0.03, p = 0.66; MPTA ρ = 0.04, p = 0.56; JLO ρ = 0.05, p = 0.39; Appendix C).

**Appendix Table B1.**
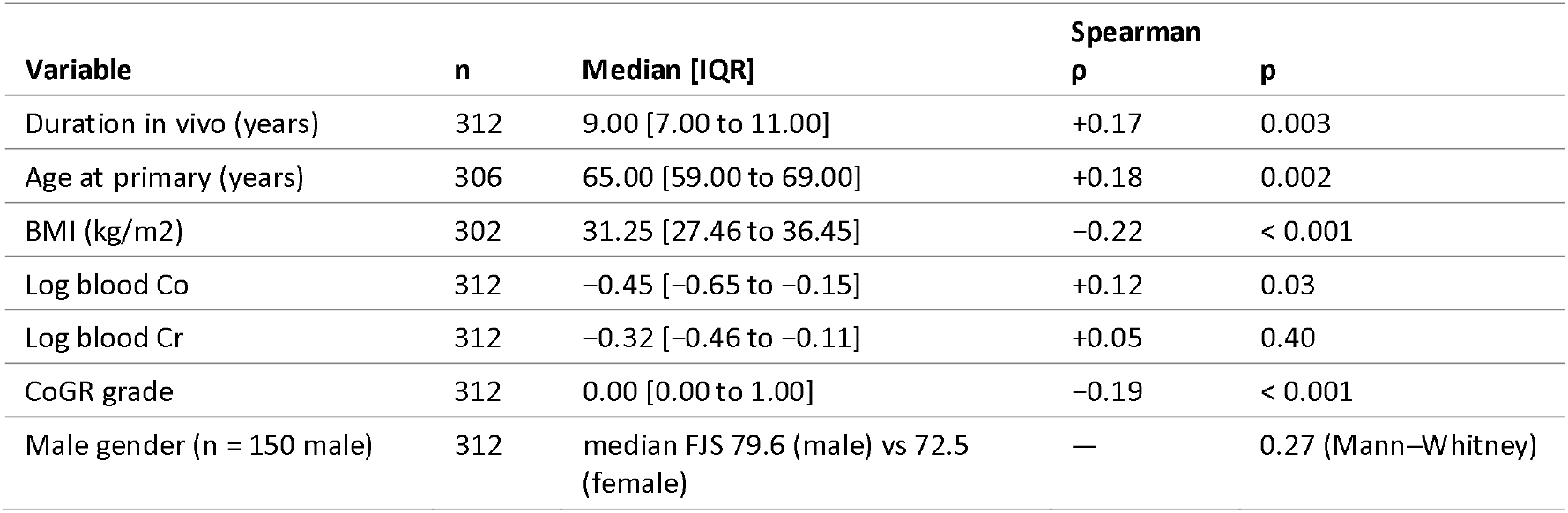
Univariable associations between FJS and candidate predictors in unrevised patients (Spearman rank correlation unless stated). Variable medians are shown with interquartile ranges; blood Co and Cr are log-transformed.

| Variable | n | Median [IQR] | Spearman $\rho$ | p |
| --- | --- | --- | --- | --- |
| Duration in vivo (years) | 312 | 9.00 [7.00 to 11.00] | +0.17 | 0.003 |
| Age at primary (years) | 306 | 65.00 [59.00 to 69.00] | +0.18 | 0.002 |
| BMI (kg/m<sup>2</sup>) | 302 | 31.25 [27.46 to 36.45] | -0.22 | < 0.001 |
| Log blood Co | 312 | -0.45 [-0.65 to -0.15] | +0.12 | 0.03 |
| Log blood Cr | 312 | -0.32 [-0.46 to -0.11] | +0.05 | 0.40 |
| CoGR grade | 312 | 0.00 [0.00 to 1.00] | -0.19 | < 0.001 |
| Male gender (n = 150 male) | 312 | median FJS 79.6 (male) vs 72.5 (female) | — | 0.27 (Mann-Whitney) |

#### B.3 Reduced model and model checks

A reduced Tobit model retaining only the variables significant in the primary model (duration in vivo, age at primary, BMI and CoGR) gave materially unchanged estimates (Appendix Table B2), indicating that the CoGR association is not an artefact of adjustment for low-information covariates. Variance inflation factors for all predictors were below 1.2, indicating no meaningful multicollinearity. Standardised residuals for the non-censored observations (n = 216) approximated a normal distribution (Shapiro–Wilk p = 0.001; skewness −0.09, excess kurtosis −0.93), the significant test result reflecting a modestly flatter-than-normal distribution rather than skew or heavy tails (Appendix Fig. B2). Leave-one-out re-estimation showed that the CoGR coefficient changed by no more than 0.94 points (<10% of its magnitude) on removal of any single patient.

**Appendix Table B2.** Full versus reduced multivariable Tobit model, FJS on candidate predictors (n = 296).

| Variable | Full model coefficient (95% CI) | Reduced model coefficient (95% CI) |
| --- | --- | --- |
| Duration in vivo (per year) | +2.81 (1.19 to 4.43) | +2.81 (1.20 to 4.42) |
| Age at primary (per year) | +1.07 (0.40 to 1.73) | +1.08 (0.42 to 1.75) |
| BMI (per kg/m<sup>2</sup>) | -0.97 (-1.76 to -0.18) | -0.96 (-1.74 to -0.17) |
| CoGR (per grade) | -9.83 (-15.44 to -4.21) | -10.22 (-15.78 to -4.67) |

**Appendix Fig. B2.**
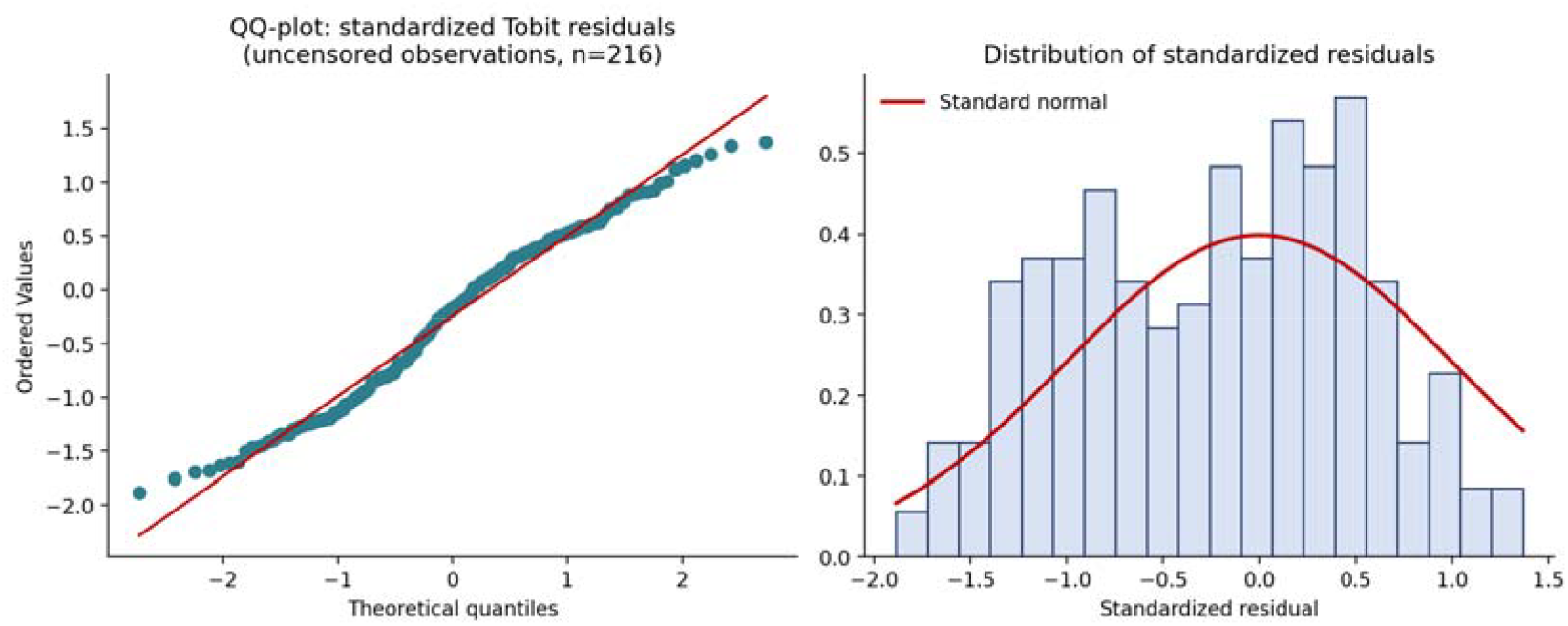
QQ-plot and distribution of standardised Tobit residuals for non-censored observations (n = 216)

#### B.4 Effect modification by cobalt

Appendix Table B3 gives the full median-split results. The finding was robust to modelling Co continuously rather than dichotomously (CoGR × log10 Co interaction −13.9, p = 0.022; likelihood-ratio p = 0.019) and to the additional inclusion of log Cr in the interaction model (interaction p = 0.014). In the high-cobalt stratum the BMI effect was stronger (−2.06 points per kg/m^2^, p < 0.001) and the duration effect weaker (+0.23 points per year, p = 0.85) than in the low-cobalt stratum (BMI −0.35, p = 0.54; duration +3.95, p < 0.001). Appendix Table B4 gives the tertile and quartile results: the CoGR effect increased stepwise across successive strata, only the highest stratum showed a clearly significant association, and individual strata are modestly powered (n = 98–99 and 74).

**Appendix Table B3.** Effect modification of the CoGR–FJS association by Co concentration (high Co defined as blood Co above the whole-sample median). Stratified and interaction Tobit models were adjusted for duration in vivo, age at primary, BMI and gender.

| CoGR effect on FJS (per grade) | n | Coefficient (95% CI) | p |
| --- | --- | --- | --- |
| Co ≤ median (stratified model) | 155 | $-4.44$ ($-12.03$ to $3.15$) | 0.25 |
| Co $>$ median (stratified model) | 141 | $-17.47$ ($-25.50$ to $-9.43$) | $< 0.001$ |
| Low-Co simple effect (interaction model) | 296 | $-3.52$ ($-10.95$ to $3.92$) | 0.35 |
| High-Co simple effect (interaction model) | 296 | $-17.29$ ($-25.66$ to $-8.92$) | $< 0.001$ |
| CoGR × high-Co interaction term | 296 | $-13.78$ ($-24.93$ to $-2.62$) | 0.015 |

**Appendix Table B4.** CoGR effect on FJS (latent points per grade) within tertiles and quartiles of blood Co (Tobit models adjusted for duration in vivo, age at primary, BMI and gender; n = 296).

| Blood Co stratum | Range (µg/L) | n | Coefficient (95% CI) | p |
| --- | --- | --- | --- | --- |
| Tertile 1 | 0.08–0.26 | 99 | –4.91 (–15.67 to 5.85) | 0.37 |
| Tertile 2 | 0.26–0.51 | 99 | –6.71 (–15.39 to 1.97) | 0.13 |
| Tertile 3 | 0.51–77.58 | 98 | –19.38 (–28.89 to –9.87) | < 0.001 |
| Quartile 1 | 0.08–0.22 | 74 | –1.52 (–13.11 to 10.07) | 0.80 |
| Quartile 2 | 0.22–0.34 | 74 | –8.88 (–19.23 to 1.48) | 0.093 |
| Quartile 3 | 0.34–0.70 | 74 | –10.63 (–21.24 to –0.02) | 0.0495 |
| Quartile 4 | 0.70–77.58 | 74 | –19.70 (–31.24 to –8.15) | < 0.001 |

#### B.5 CoGR as grade groups and ordinary regression

To check the assumption of a straight-line relationship, CoGR was grouped as < 0 (n = 29), 0 (n = 148; reference), 1 (n = 83) and ≥ 2 (n = 36) in a Tobit model adjusted as in the primary model. Relative to grade 0, FJS was higher in the < 0 group (+15.9 points, p = 0.11) and lower at grade 1 (−9.5, p = 0.11) and grade ≥ 2 (−19.6, p = 0.016); the estimates fall in the same order as the linear trend although only the last reached significance. An OLS model identified the same four significant predictors with the same directions of effect (duration +2.19, p = 0.001; age +0.79, p = 0.002; BMI −0.67, p = 0.043; CoGR −6.98, p = 0.003; adjusted R2 = 0.11).

#### B.6 CoGR in each case–control group

CoGR rose in step with disease severity (Appendix Table B5; all case–control comparisons p < 0.001, Mann–Whitney). A CoGR grade of 1 or more was present in 11 of 12 ALVAL cases (92%) versus 31% of controls (Fisher’s exact odds ratio 24.4, p < 0.001); given the sample size this odds ratio carries very wide uncertainty and the per-grade regression estimates should be regarded as the primary effect sizes. Among revised patients, CoGR ≥ 1 was present in 19 of 28 ARMD cases and 11 of 12 ALVAL cases but in only 2 of 19 without ARMD (both p < 0.001), and CoGR distributions differed (Mann–Whitney: ARMD vs histology-negative p < 0.001; ALVAL vs histology-negative p < 0.001).

**Appendix Table B5.** CoGR grade in each group. CoGR rose in step with disease severity (all case– control comparisons p < 0.001, Mann–Whitney).

| Group | n | Median CoGR [IQR] | CoGR ≥ 1 |
| --- | --- | --- | --- |
| Controls — unrevised, FJS ≥ 75 | 158 | 0 [0 to 1] | 53 (34%) |
| Controls — revised, histology negative | 19 | 0 [–0.5 to 0] | 2 (11%) |
| ARMD (histology confirmed) | 28 | 1 [0 to 2] | 19 (68%) |
| ALVAL subset | 12 | 2 [1 to 2.25] | 11 (92%) |

#### B.7 Sensitivity of the odds ratios to the estimation method

Odds ratios were closely similar under maximum likelihood, Firth, FLIC and FLAC (Appendix Table B6), with FLAC giving the most conservative estimates. Mean predicted probabilities under Firth regression slightly exceeded the observed event proportions (for example 7.0% versus 6.5% in the adjusted ALVAL model), as expected, and were restored by FLIC and FLAC. Maximum-likelihood estimates were finite in every model, indicating no separation. When a quadratic term for CoGR was added there was weak evidence of flattening of the log-odds at the highest grades (penalised likelihood-ratio p = 0.037– 0.047, with only 3–5 patients at grades 3 and 4).

**Appendix Table B6.** Odds ratio per one-grade increase in CoGR (95% CI) under maximum likelihood (ML), Firth, Firth with intercept correction (FLIC) and Firth with added covariate (FLAC). FLIC slopes equal Firth’s by construction (only the intercept is re-estimated), so the Firth interval applies. FLAC intervals are profile-likelihood intervals from the augmented-data fit (approximate). Events per covariate (EPV) shown for each model; mean predicted probability is compared with the observed event proportion.

| Model | Cases /<br>n | EPV | ML | Firth | FLIC | FLAC |
| --- | --- | --- | --- | --- | --- | --- |
| ARMD unadjusted | 28 / 205 | 28.0 | 1.92 (1.34–2.77) | 1.90 (1.34–2.75) | 1.90 (1.34–2.75) | 1.88 (1.32–2.72) |
| ARMD adjusted | 27 / 196 | 5.4 | 2.05 (1.36–3.09) | 1.97 (1.34–2.98) | 1.97 (1.34–2.98) | 1.93 (1.32–2.89) |
| ALVAL unadjusted | 12 / 189 | 12.0 | 3.19 (1.87–5.45) | 3.05 (1.89–5.34) | 3.05 (1.89–5.34) | 2.97 (1.84–5.17) |
| ALVAL adjusted | 12 / 186 | 4.0 | 3.39 (1.91–6.03) | 3.12 (1.89–5.62) | 3.12 (1.89–5.62) | 2.98 (1.82–5.28) |

### Appendix C. Radiographic measures

#### C.1 Radiographic measures and the CoGR models

Radiographic measures were available for 260 of the 312 unrevised patients (83%), whose age, BMI, CoGR and FJS were similar to the full unrevised cohort. The lateral distal femoral angle (LDFA), the medial proximal tibial angle (MPTA) and joint line obliquity (JLO) are summarised in Appendix Table C1. As JLO is the sum of LDFA and MPTA, it was examined in a separate model rather than alongside them. None of the three measures were associated with FJS (Spearman ρ 0.03 to 0.05; all p ≥ 0.39) or with CoGR (all p ≥ 0.34). Adding LDFA and MPTA to the primary Tobit model did not change the CoGR estimate (−9.6 points per grade with and without them; likelihood-ratio test for the two angles p = 0.43), and adding JLO gave the same result (−9.7). The CoGR × cobalt interaction was also unchanged (−17.3 points, p = 0.006, with and without radiographic adjustment), as was the CoGR effect in patients with above-median cobalt (−18.7 without, −18.4 with LDFA and MPTA, −18.8 with JLO). Alignment therefore did not explain the association between CoGR and FJS.

**Appendix Table C1.**
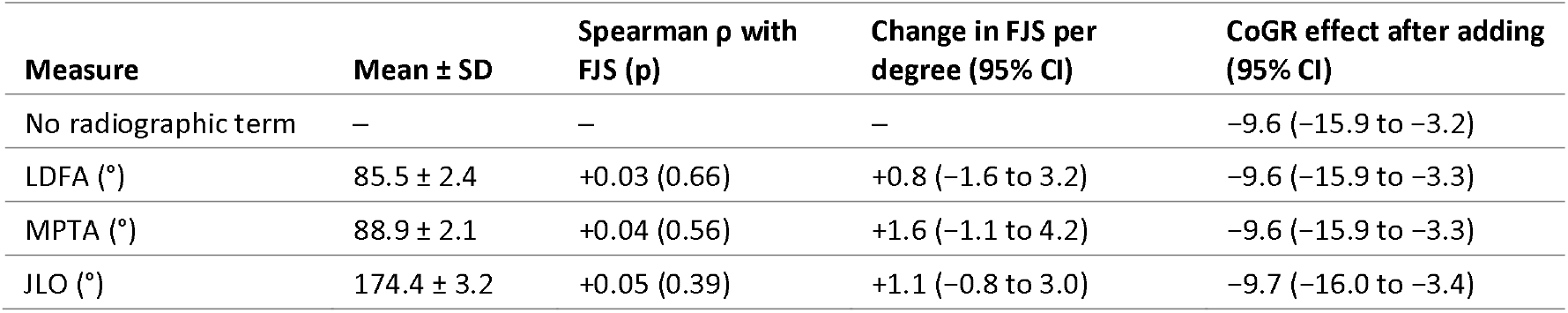
Radiographic measures in unrevised patients (n = 260), their association with FJS, and the effect of adding each to the primary Tobit model. Models were adjusted for duration in vivo, age at primary, BMI, gender and log Cr (n = 246 complete cases). LDFA and MPTA were entered together; JLO, being their sum, was entered in a separate model.

| Measure | Mean ± SD | Spearman $\rho$ with FJS (p) | Change in FJS per degree (95% CI) | CoGR effect after adding (95% CI) |
| --- | --- | --- | --- | --- |
| No radiographic term | – | – | – | -9.6 (-15.9 to -3.2) |
| LDFA (°) | 85.5 ± 2.4 | +0.03 (0.66) | +0.8 (-1.6 to 3.2) | -9.6 (-15.9 to -3.3) |
| MPTA (°) | 88.9 ± 2.1 | +0.04 (0.56) | +1.6 (-1.1 to 4.2) | -9.6 (-15.9 to -3.3) |
| JLO (°) | 174.4 ± 3.2 | +0.05 (0.39) | +1.1 (-0.8 to 3.0) | -9.7 (-16.0 to -3.4) |

